# Preparedness, Perceived Impact and Concerns of health Care Workers in a Teaching Hospital during Coronavirus Disease 2019 (COVID-19)

**DOI:** 10.1101/2020.07.15.20140095

**Authors:** Kumar Saurabh, Shilpi Ranjan

**Affiliations:** Department of Pediatrics, Government Medical College, Bettiah, Bihar, India., Email id; Department of Community Medicine, Nalanda Medical College and Hospital, Patna, Bihar, India., Email id

**Keywords:** COVID-19, HCW, Pandemic, Preparedness, Anxiety, Stress, Hydroxychloroquine

## Abstract

**Objective:** Coronavirus Disease 2019 is a new threat to human lives worldwide. Preparedness of institutions during epidemic outbreak has a pivotal role in saving lives and preventing further spread. At the same time, these pandemics impact badly on professional and personal life of Health care workers. The objective of this study is to find the opinion of Health care workers regarding their level of preparedness, concerns and perceived impact related to this pandemic outbreak.

**Materials and Methods:** in this study, random samples of doctors and nurses was provided with a self-administered questionnaire regarding their preparedness, work and non-work related concerns and impact on their lives during Covid-19 outbreak.

**Results:** Most of the Health Care Workers believed that their institute preparation to fight Covid-19 pandemic is better than prior to onset of this crisis (p⍰0.001). Work related stress was seen more commonly in nurses whereas higher frequency of non-work related stress was observed among doctors. Nurses (75.55%) faith in their employer was more than doctors faith (46.66%) regarding their medical needs. There was more acceptance of hydroxychloroquine as a prophylactic drug for Covid-19 in doctors compared to nurses (p ⍰0.01).

**Conclusions:** Though this institute was more prepared at the time of pandemic spread, substantial opportunity of improvement remains. The consistency of work and non work related anxiety and stress in health care workers is very high in present study group. Concerns and risks of Health Care Workers should be addressed ethically and adequately by strengthening safety measures and building trust in the system they work.

## Introduction

In December 2019, a series of pneumonia cases of unknown origin were identified in Wuhan, the capital city of Hubei province, China with features similar to viral pneumonia ^[1]^. The pathogen has been identified as a novel enveloped RNA beta coronavirus. The World Health Organisation (WHO) officially named the disease as coronavirus disease 2019 (COVID-19) and the pathogen as severe acute respiratory syndrome coronavirus 2 (SARS-Cov-2) on 11^th^ February 2020^[2]^. Person to person transmission has been documented as patients with the infection have been identified both in hospitals and in family settings ^[3],[4]^

The World Health Organisation (WHO) declares Coronavirus disease 2019 (COVID-19) as a pandemic on 11^th^ March 2020^[5]^. As on 18 April 2020, globally laboratory confirmed cases were 2160207 with 146088 deaths ^[6]^. As per government of India record 13295 active cases of Covid-19 including Health Care Workers (HCW) were identified in India with 519 deaths and 2301 patients cured ^[7]^. Most of the worst affected countries including India started complete lockdown of nation on 25^th^ March 2020^[8]^.

The ability to respond quickly in the face of an emerging novel coronavirus disease 2019 is critical for safety of patients and their near ones. At this point of time, the capacity to work together across institutions and countries has proved most important step in the control of recent pandemics such as COVID-19. Indeed, these situations provide an opportunity to help in policy making in real time. ^[9],[10]^

Health Care Workers (HCW) are at higher risk themselves of contracting COVID-19, and thus they can place their patients at risk. In China, more than 3300 HCW have been infected and at least 22 had died. In Italy, almost 20% of HCW involved in COVID-19 treatment were infected, and some have died.^[11]^ Remember, unlike ventilators or wards HCW cannot be urgently manufactured or run at 100% occupancy for long periods. In this context government should treat HCW as human individuals and not simply as pawns to be deployed.

We therefore investigate the preparedness, concerns and perceived impacts of doctors and nurses (HCW) at a medical college of north India.

## Materials &Methods

A random sample of doctors and nurses of Government Medical College, Bettiah, India was invited to participate in the study during initial days of disease outbreak in India (From 1 March to 31 March2020). The admission of first batch of medical students to this institute was started in 2013. This is a tertiary care teaching hospital which could serve over four million populations. At this point of time two hundred bed corona isolation wards has been created so that patients with this disease could seek medical care.

Study was approved by institutional ethics committee. Written informed consent was taken from every participant after explaining them objective of the study. All participants were provided with a preformed self-administered questionnaire which includes different sections in order to investigate their demographic data, education, years of experiences, PPE training, preparedness, concerns and perceived impact. Our questionnaire was adopted from a previous study of Singapore and Egypt ^[12,13]^. Sections were as follows.

1. Demographic data.
2. Preparedness of doctors and nurses for Covid-19 pandemic.
3. Work related concerns during Covid-19 pandemic.
4. Non-work related concerns during Covid-19.
5. Perceived impacts of HCWs.

In statistical methods, first descriptive analysis was done which includes frequency, percent, mean, standard deviation. Pearson’s Chi square test for categorical variables was used to compare doctors and nurses. Level of significance was set at p⍰ 0.05. Data entry and statistical analysis were performed using the Statistical Package for Social Science (SPSS) version 26.0.

## Results

There were 120 respondents (75 doctors and 45 nurses), in which 53 (44.11%) were females. The age distribution of respondents was as follows: 20-29 years, 16 (13.33%); 30-39 years, 61 (50.83%); 40-49 years, 32 (26.66%); 50-59 years, 9 (7.5%); 60-69 years, 2 (1.66%). The participant doctors were consultants (10.66%), Assistant Professor/Assistant consultants (36.00), rest were residents (53.33%). Experience of respondents in the field of health care was as follows; ⍰5 year, 13 (10.83%); 5-10 years, 62 (51.66); 11-15 years, 27 (22.50); ⍰15 years, 18 (15.00).

Most of the HCWs agree that their institute was not ready appropriately to respond to this crisis. As only 18.66% of doctors and 17.77% of nurses believed that their institute was ready prior to this pandemic. But in contrast, when asked about preparedness at this point, most of nurses (60%) answered yes against 37.33% of doctors with a significant p-value of 0.015. when we compared answers of all 120 respondents prior to and at real time in terms of preparedness, we found a statistically significant difference of p⍰0.001. It means this institute has done well in this available time frame of ‘prior’ and ‘at this point’ [Table 1]. Second important result of table 1 is acceptance of hydroxychloroquine as a prophylactic drug for Covid-19 in doctors compared to nurses (p ⍰0.01). One more point of relevance is that 56% of doctors and 40% of nurses noticed that during Covid-19 crisis other infection prevention related activities were neglected.

**Table 1.** Preparedness of Health Care Workers for Covid-19 pandemic.

| **Preparedness for Covid-19 Pandemic** | **Doctors**<br>**N=75 (%)** | **Nurses**<br>**N= 45(%)** | **P value** |
| --- | --- | --- | --- |
| Covid-19 is a an important health problem. | 74 (98.66) | 42 (93.33) | 0.11 |
| Covid-19 is more important than the other infection control-related problems that I deal with on a regular basis. | 56 (74.66) | 40 (88.88) | 0.06 |
| Prior to Covid-19 emergence, locally endemic diseases were viewed as an important health problem at my institution. | 39 (52.00) | 24 (54.33) | 0.88 |
| Senior-level hospital administrators believed that Covid-19 was a more important problem than I thought it was. | 63(84.00) | 39 (86.66) | 0.69 |
| Prior to the current Covid-19 crisis, my hospital was well prepared for a potentially pandemic situation such as Covid-19. | 14 (18.66) | 8 (17.77) | 0.90 |
| At this point in time, my hospital is well prepared for a potentially pandemic situation such as Covid-19. | 28 (37.33) | 27 (60.00) | 0.015 |
| During the Covid-19 crisis, adequate resources to respond to the crisis were provided. | 27 (36.00) | 11 (24.44) | 0.18 |
| During the Covid-19 crisis, other important infection prevention-related activities were neglected. | 42 (56.00) | 18 (40.00) | 0.08 |
| N-95 masks have been readily available throughout the Covid-19 crisis at my institution. | 5 (6.66) | 3 (6.66) | 1.0 |
| The recommendation for airborne precautions for suspected Covid-19 cases was appropriate at the beginning of the Covid-19 | 28 (37.33) | 17 (37.77) | 0.96 |
| crisis at my institution. |  |  |  |
| Surgical masks have been readily available throughout the Covid-19 crisis at my institution. | 7 (9.33) | 4 (8.88) | 0.93 |
| Alcohol based hand hygiene products have been readily available throughout the Covid-19 crisis at my institution. | 9 (12.00) | 3 (6.66) | 0.34 |
| All healthcare workers should be mandated to receive the prophylactic dose of Hydroxychloroquine. | 56 (74.66) | 23 (51.11) | 0.008 |

Table 2 shows 88.88% nurses in comparison of 65.33% doctors agree that risk of contracting Covid-19 is a part of their job (p⍰0.01). It is surprising that fewer doctors rely on their employer than nurses when it comes to medical needs with a statistically significant difference (p⍰0.01). It is good to find that most of the respondents either don’t consider resigning or considering it acceptable for their colleagues.

**Table 2:** Work related concerns of Health Professional due to Covid-19.

| **Work related concerns**<br>**Of Health Professional** | **Doctors**<br>**N= 75 (%)** | **Nurses**<br>**N= 45 (%)** | **P value** |
| --- | --- | --- | --- |
| My job would put me at great risk of exposure to Covid-19 | 71 (94.66) | 44 ( 97.77) | 0.39 |
| I am afraid of falling ill with covid-19 | 66 (88.00) | 36 (80.00) | 0.23 |
| I feel that I should not be looking after patients with Covid-19 | 21 (28.00) | 18 (40.00) | 0.17 |
| I would accept that the risk of contracting Covid-19 is part of my job | 49 (65.33) | 40 (88.88) | 0.0043 |
| I might look for another job or consider resigning because of the risk of contracting Covid-19 | 6 (8.00) | 5 (11.11) | 0.56 |
| I would consider it acceptable if my colleagues resign because of their fear of Covid-19 | 7 (9.33) | 6 (13.33) | 0.49 |
| I am confident that my employer would look after my medical needs if I were to fall ill with | 35 (46.66) | 34 (75.55) | 0.0019 |
| Covid-19 |  |  |  |

There is higher non work related concerns in doctors than nurses. About 92% of doctors have higher worries of spreading infection to their families compared to 64.44% of nurses (p⍰0.01). Higher number of doctors (97.33%) also believed that people close to them would be worried as they may get infected by them compared to nurses with a p value of ⍰0.01 [Table 3].

**Table 3:** Non-work related concerns of Health Professional due to Covid-19.

| **Non-work related concerns of Health Professional** | **Doctors**<br>**N= 75 (%)** | **Nurses**<br>**N= 45 (%)** | **P value** |
| --- | --- | --- | --- |
| people close to me would be at high risk of getting Covid-19 because of my job | 69 (92.00) | 29 (64.44) | 0.00016 |
| I would be concerned of my family. | 70 (93.33) | 41 (91.11) | 0.65 |
| I would be concerned of my friends. | 66 (88.00) | 40 (88.88) | 0.88 |
| I would be concerned of my colleagues. | 66 (88.00) | 41 (91.11) | 0.59 |
| people close to me would be worried for my health | 70 (93.33) | 37 (82.22) | 0.06 |
| people close to me would be worried as they may get infected by me. | 73 (97.33) | 35 (77.77) | 0.0005 |

This study also observes that Covid-19 has a higher impact on personal life and work of doctors than nurses [Table 4]. Doctors feel more stressed and assume that they would have to do work more and have to do the work not normally done by them (p⍰0.01).

**Table 4:** Perceived impact on personal life and work of Health Professional due to Covid-19.

| **Perceived impact on personal life and work of Health Professional** | **Doctors**<br>**N= 75 (%)** | **Nurses**<br>**N= (%)** | **P value** |
| --- | --- | --- | --- |
| I would be afraid of telling my family about the risk I am exposed to | 42 (56.00) | 28 (62.22) | 0.50 |
| people would avoid me because of my job | 33 (44.00) | 26 (57.77) | 0.14 |
| people would avoid my family members because of my job | 39 (52.00) | 24 (53.33) | 0.88 |
| I would avoid telling other people about the nature of my job | 21 (28.00) | 13 (28.88) | 0.91 |
| there would be adequate staff at my workplace to handle the increased demand | 36 (48.00) | 18 (40.00) | 0.39 |
| there would be more conflict amongst colleagues | 56 (74.66) | 26 (57.77) | 0.54 |
| at work |  |  |  |
| I would feel more stressed at work | 71 (94.66) | 33 (73.33) | 0.0008 |
| I would have an increase in workload | 56 (74.66) | 32 (71.11) | 0.66 |
| I would have to work overtime | 55 (73.33) | 12 (26.66) | 0.00001 |
| I would have to do work not normally done by me | 71 (94.66) | 16 (35.55) | 0.00001 |

## Discussion

Adequacy and appropriateness of institutional preparedness and response to emergency situation like emerging and re-emerging pandemics is a key to safety of mankind. In this regard, response and experience from on-going crisis will be very helpful for future interventions. Result of this study provides a well oriented and timely perspective on the response to the current Covid-19 crisis. This tertiary care teaching institute plays a vital front line role in the management of a large population. Improvement in present scenario will be helpful in saving many lives. As per the mandate of reorientation of medical education scheme, this medical college is also linked to primary health facilities in rural and urban area. So primary care physicians will be also affected by the measures taken by this institute.^[14]^

In this study, we found that majority of HCW felt that their institute was not well prepared for COVID-19 crisis and was not provided with adequate resources. HCW did not receive any training regarding use of PPE (personal protection equipment) or any other infection control related training before the onset of this crisis. But with the progression of disease burden, many HCWs including doctors and nurses received training for PPE use through online courses conducted by government of India. Accessibility of HCWs to PPE, N-95 mask, surgical mask and alcohol based hand hygiene products in the events of pandemic is a critical component of preparedness plan. In this respect, it is very concerning that most doctors and nurses reported a shortage of these products. HCWs in the other parts of world are also working in the same scenario with inadequate resources.^[11],[12]^ world Health Organisation has also discussed this issue in a guideline stating that there is shortage of personal protective equipment in many parts of world which is endangering health workers lives.^[15]^

In view of severe acute respiratory syndrome and the threat of bioterrorism has lead to heroic efforts to address pandemic preparedness at health care institutions throughout the country. It is very much discouraging that the majority of respondents 64% doctors and 75.56% nurses believed that they do not have adequate resources to deal with Covid-19 pandemic. However many HCWs accept that, though their institution was not well prepare prior to the pandemic but with progression of pandemic, scenario improved and institute was in a better condition to tackle this acute crisis (p⍰0.001).

This medical college has faced its first pandemic and this Covid-19 crisis represents the first real test of preparedness. However, there is considerable opportunity for improvement remains as results of this study suggest. More effort is required to further refine and improve preparedness plans. We should plan to prepare in such a way that other vital health care activities and other infection control related activities should not be ignored during emergence of a new infection such as Covid-19 that may last for several months. Many of the participants of this study group (56% doctors and 40% nurses) accepted that other infection control related activities were neglected during this period.One recent study from India also concluded that other vital health care activities like infection control, immunisation services and maternal care services were compromised due to COVID-19 pandemic. ^[14]^

Our results match with the outcome of previous study conducted in Germany and Egypt, where majority of HCW recognised the obligation to treat patients despite high risk.^[13,16]^ It is well known that during an outbreak, HCW are expected to work for longer period under high pressure and often with inadequate resources. At the same time they have to accept risks associated with close contact of severely ill patients. This necessarily increases their anxiety level and concerns.

Ministry of health and family welfare, has recommended chemoprophylaxis with hydroxychloroquine (400 mg twice on day 1, then 400mg once a week thereafter) for asymptomatic HCWs treating patients with suspected or confirmed Covid-19 cases.^[17]^ Only 51% of nurses took the doses as compared to 74.66% of doctors. Prolongation of QT interval, its interaction with Azithromycin and glucose −6-phophate dehydrogenase deficiency were different concerns associated with this medicine in HCW. However, the use of hydroxychloroquine at large scale is of concern because the drug is untested, the benefits unknown and the risks not negligible.^[18]^

From our results it is clear that there are higher non work related stress and family stress among physician compared to nurses ^[13,16]^. Result of this study coincides with previous studies. Around 50% of doctors and nurses admitted that people will avoid them and their family members because of nature of their job. Previous studies conducted at Singapore and Egypt show similar results [12,13]. Doctors were more worried than nurses about their stress level, overtime and extra workload.

This study highlights the contradiction between how HCW might feel about their professional duty for their patients and their personal duty to themselves and their families. Alongside concerns for their personal safety, HCW are anxious about passing infections to their families.^[19]^ The level of commitment of HCWs during a pandemic is potentially affected by these issues. This should be addressed in any national pandemic preparedness plan.^[20]^

Outbreaks like Covid-19 are critical reminders of the significance of public health readiness and the need for continued strengthening of public health agencies. This would be possible only by prior investments in public health preparedness. Moreover, problems associated with nationwide lockdown, migration of workers from big cities to their native place, psychological impact of quarantine, supply of daily needs like food and water, and financial support are other relevant points to be addressed for pandemic preparedness.^[21]^

Before reaching any conclusion based on this study, it is necessary to consider that this study had several potential limitations. Here, self-administered questionnaire was a method to investigate. Response rate is typically low in these types of questionnaire, also characteristics and view of other HCW is not known. Because of limited geographical limitations, study findings may not be generalised. But this study represents only a very few and early studies from India about the preparedness and concerns of HCW in response to COVID-19 pandemic.

In conclusion, this study provides an important insight into the concerns, perceived impact and preparedness of health care workers of an institute. Substantial revision of pandemic preparedness plans is required to improve the present scenario on the basis of current experience. To ensure effective functioning of an institute during public health emergencies government should considerably invest in infrastructure, capacity building and strengthening of health care services. The consistency of work and non work related anxiety and stress in health care workers is very high in present study group. So, professional ethical guidelines allowing for balancing the needs of society and personal risks are required to help HCW to fulfil their duties.

## Data Availability

all data is included in the uploaded article

## Acknowledgements

The authors would like to thank all healthcare workers who participated in our study and for being as frontline warrior against Covid-19.

